# A Novel Olfactory Self-Test Effectively Screens for COVID-19

**DOI:** 10.1101/2021.02.18.21251422

**Authors:** Kobi Snitz, Danielle Honigstein, Reut Weissgross, Aharon Ravia, Eva Mishor, Ofer Perl, Shiri Karagach, Abebe Medhanie, Nir Harel, Sagit Shushan, Yehudah Roth, Behzad Iravani, Artin Arshamian, Gernot Ernst, Masako Okamoto, Cindy Poo, Niccolò Bonacchi, Zachary Mainen, Erminio Monteleone, Caterina Dinnella, Sara Spinelli, Franklin Mariño-Sánchez, Camille Ferdenzi, Monique Smeets, Kazushige Touhara, Moustafa Bensafi, Thomas Hummel, Johan N. Lundström, Noam Sobel

## Abstract

Key to curtailing the COVID-19 pandemic are wide-scale testing strategies^1,2^. An ideal test is one that would not rely on transporting, distributing, and collecting physical specimens. Given the olfactory impairment associated with COVID-19^3-7^, we developed a novel measure of olfactory perception that relies on smelling household odorants and rating them online. We tested the performance of this real-time tool in 12,020 participants from 134 countries who provided 171,500 perceptual ratings of 60 different household odorants. We observed that olfactory ratings were indicative of COVID-19 status in a country, significantly correlating with national infection rates over time. More importantly, we observed remarkable indicative power at the individual level (90% sensitivity and 80% specificity). Critically, olfactory testing remained highly effective in participants with COVID-19 but without symptoms, and in participants with symptoms but without COVID-19. In this, the current odorant-based olfactory test stands apart from symptom-checkers (including olfactory symptom-checkers)^3^, and even from antigen tests^8^, to potentially provide a first line of screening that can help halt disease progression at the population level.

## RESULTS

### Olfactory perception indicates on levels of COVID-19 infection at the population level

We built an online tool (www.smelltracker.org) where each participant selected 5 of 71 possible household odorants (Extended Data Table 1), and then smelled and rated each using visual-analogue scales for perceived intensity and pleasantness, namely the primary dimensions of olfactory perception^9^. Participants also completed a common symptom-check (reporting on fever, cough, subjective loss of taste and smell, etc), and reported on any formal COVID-19 testing they had undergone. Participants could generate a user-name to track their own performance over time, but the tool was otherwise completely anonymous to protect user privacy^10^.

Between the dates of March 25^th^ 2020 and September 23^rd^ 2020, we collected data from 12,020 individuals (7,189 Women, mean age = 44.32 ± 14.28, 4,831 Men, mean age = 45.23 ± 15.29) (Extended Data Figure 1A), residing in 134 countries (Extended Data Figure 1B, 1C). Of these, 348 participants reported positive COVID-19 test results (C19+), 400 participants reported negative test results (C19-), and the COVID-19 status of the remaining 11,272 participants was unknown (C19-UD) (Extended Data Figure 1D). The presence of at least one disease symptom was reported by 91.1% of C19+ participants, 55.3% of C19-participants, and 55% of C19-UD participants (Extended Data Figure 1E). Participants could use the online tool repeatedly, yet 10,103 participants (84%) used it only once, 1,130 participants (9.4%) used it twice, and the remaining 6.6% of participants used it various number of times (Extended Data Figure 1F). In total, the 12,020 participants provided 171,500 ratings applied to 60 different odorants (i.e., 11 odorants were never rated) (The entire raw data file is available in Supplementary Data File 1).

To probe for any gross differences in olfactory perception between C19+, C19-, and C19-UD, we plotted their overall odorant intensity and pleasantness estimations (Figure 1A, 1B, 1C). Consistent with previous reports^3-7^, these gross plots revealed pronounced differences between groups in intensity (two-sided Kolmogorov-Smirnov, D = 0.45, p = 3.96e-233, corrected) and pleasantness (D = 0.31, p = 2.36e-114, corrected) (Figure 1A, 1B, 1C). To provide a finer-grain view of this, we examined individual odorants, restricting our analysis to odorants that were rated by at least 25 C19+, thus retaining 23 odorants. Because a Kolmogorov-Smirnov normality of distribution test revealed that data for some of the odorants was not normally distributed, we proceeded with a non-parametric approach. A Chi Squared test comparing C19+ and C19-ratings was significant for each of the 23 odorants (all Chi Square > 7.6, all p < 0.0058, all Eta squared effect size > 0.08) (Figure 1D), and the same test on pleasantness ratings was significant for 17 of the 23 odorants (all Chi Square > 8.69, all p < 0.0188, all Eta squared effect size > 0.02) (Figure 1E) (we note that replicating this analysis using a parametric analysis of variance yielded nearly identical results).

**Figure 1.**
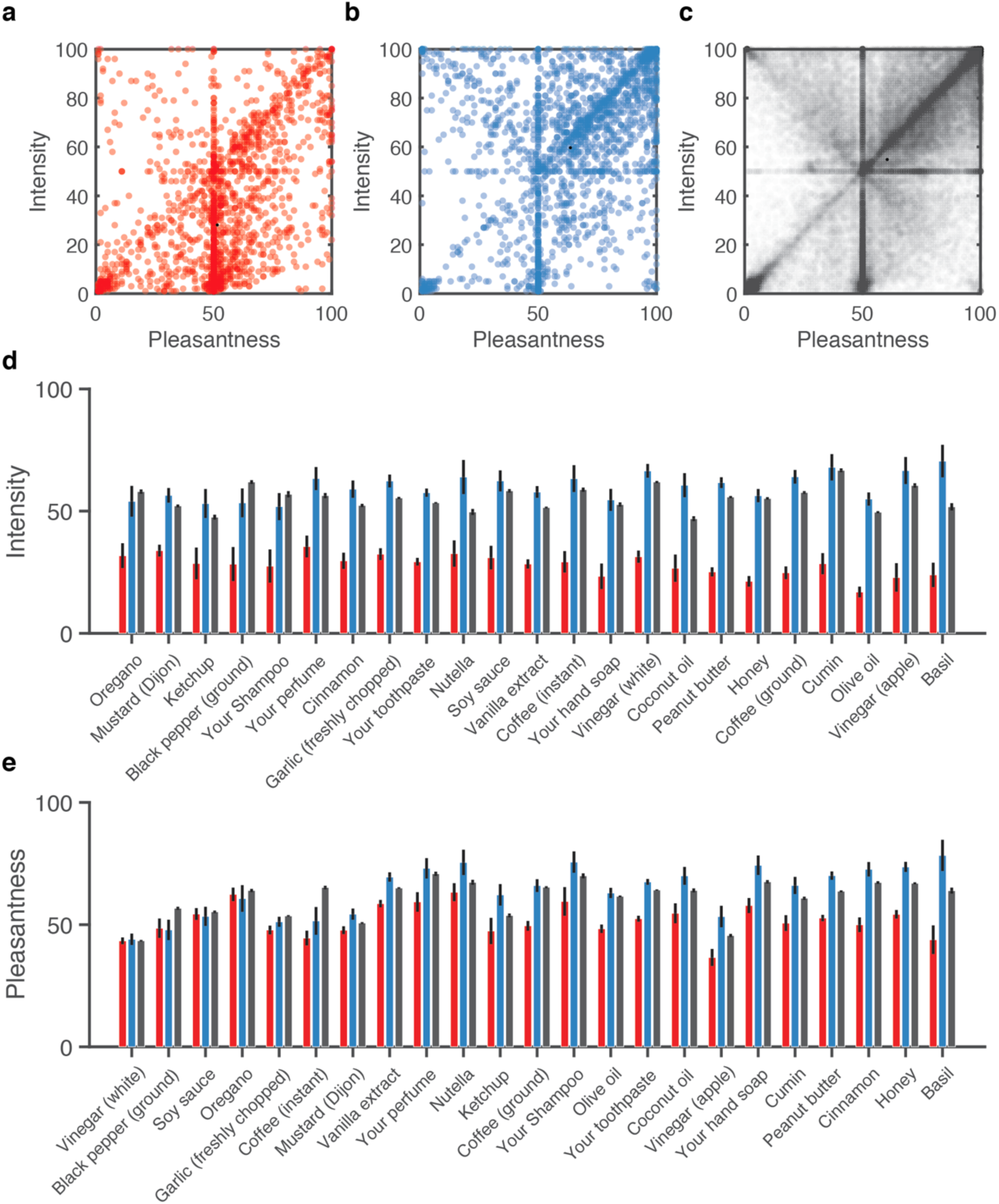
Olfactory perception indicates on levels of COVID-19 infection at the population level. For A-C, each dot is an odorant rating, aligned for its pleasantness and intensity estimates: **A**. All ratings from C19+ participants (n = 2670 ratings). **B**. All ratings from C19-participants (n = 2580 ratings). **C**. All ratings from C19-UD participants (n = 80,500 ratings). **D**. Intensity estimates for the 23 odorants that were each rated by at least 25 C19+ participants, ordered by effect-size from low (left) to high (right). **E**. Pleasantness estimates for the 23 odorants that were each rated by at least 25 C19+ participants, ordered by effect-size from low (left) to high (right). C19+ in red, C19-in blue, and C19-UD in black. Error bars are SEM.

Having observed that, consistent with previous reports^3-7^, these gross measures of perception implied altered olfaction in COVID-19, we asked whether they were related to the COVID-19 status over time in the different countries where we collected data. Consistent with an initial analysis^11^, we concentrated on odorant intensity estimates in this global analysis, and limited this to countries with at least 250 respondents of which at least 10 were formally diagnosed. This limited us to 8 countries, where country-specific rates of COVID-19 infection over time were obtained from The Johns Hopkins University Coronavirus Resource Center^12^. We observed a significant relationship between overall group-level odorant intensity ratings and daily rates of COVID-19. More specifically, mean intensity ratings and COVID-19 prevalence were significantly correlated in 7 out of 8 countries (Sorted by Pearson correlation FDR corrected: Israel: n = 2734, r = 0.68, p < 0.0001; Sweden: n = 6133, r = 0.55, p < 0.0001; Portugal: n = 685, r = 0.47, p < 0.0001; Brazil: n = 764, r = 0.39, p < 0.0001; UK: n = 290, r = 0.26, p = 0.0011; Japan n = 290, r = 0.25, p = 0.0011; USA: n = 2276, r = 0.25, p = 0.0011; France: n = 655, r = −0.14, p = 0.09) (Figure 2). Although the country-specific sample sizes are not overwhelming in this sub-analysis, these results imply that olfactory testing can augment symptom-tracking^13^ to aid country-level rapid policy decisions related to the spread of COVID-19.

**Figure 2.**
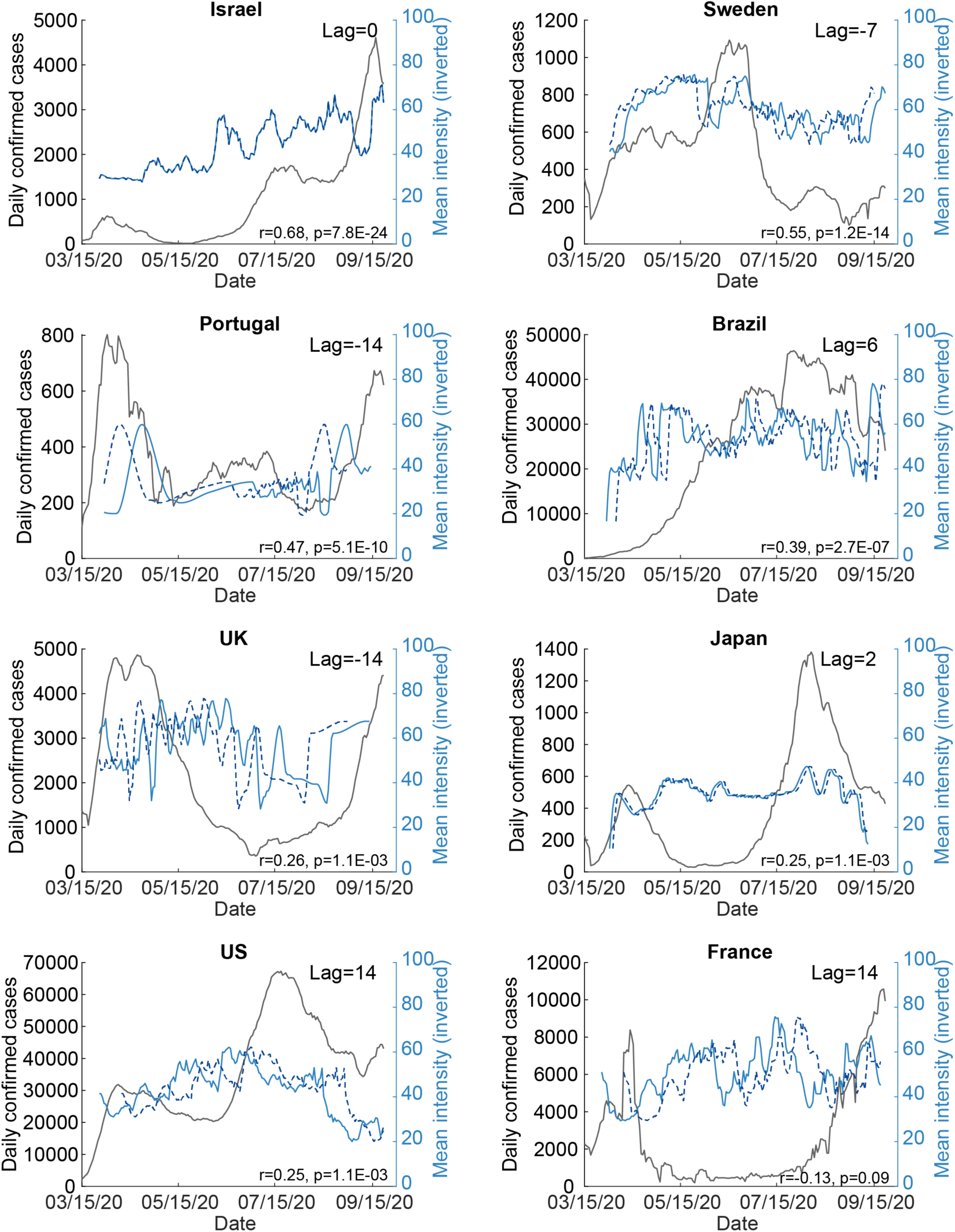
Odorant intensity estimates correlated with national COVID-19 infection rates over time. Each panel depicts the correlation between intensity estimates and national levels of COVID-19 for the given country noted at the top of the panel. Blue line: mean daily additive inverse intensity ratings. Dashed blue line: shifted additive inverse intensity time-series, after finding the peak lag using cross correlation (see Methods). Black line: number of daily confirmed cases in each country. Note that when the lag is close to zero, then the dashed and solid blue lines align and overlap.

### Olfactory perception indicates on COVID-19 at the individual level

Having observed that olfaction provides for an indication on levels of COVID-19 infection at the country level, we next asked whether it can provide an indication on COVID-19 in an individual. Given the primacy of intensity estimates at the populational level, we initially concentrated on those. We continued with the 23 odorants that were rated by at least 25 C19+ participants. These ranged in usage from the odorant ‘black pepper ground’ that had the smallest number of positively diagnosed raters at only 26 C19+ and 43 C19-participants, to the odorant ‘your toothpaste’ that was rated by as much as 336 C19+ and 330 C19-participants. We then plotted receiver operator curves (ROCs) to gauge the potential classification power of intensity estimates associated with each of these odorants. We observed remarkable classification potential, with 7 odorants generating ROCs with area under the curve (AUC) greater than 0.8 (Figure 3a, 3b). Most remarkable was the odorant Basil, with an AUC of 0.91. ROC classification success can be estimated at different true positive (sensitivity) rates selected by the observer. For context on sensitivity, we consider antigen tests, as these are promoted as a cheaper and more available alternative to RT-PCR, and have been directly compared to the latter. Antigen tests have obtained sensitivity of 30.2% in direct comparison to RT-PCR^8^ (although better results have been reported^14^, they have also been questioned^15^). In the case of the above Basil-derived ROC, at a true positive rate of 62% (namely double that of antigen tests, and on par with lower-bound estimates for RT-PCR itself^16^), we retain a remarkably low 5% false positive rate, translating to 62% sensitivity and 95% specificity (95% confidence on sensitivity: 43%-80%, 95% confidence on specificity: 75%-100%, p = 0.03 corrected, PPV = 0.94, NPV = 0.65, Matthews Correlation Coefficient = 0.58) in detecting COVID-19 using intensity estimates of the odorant Basil alone. In turn, at a more conservative true positive rate of 79%, we retain a modest 13% false positive rate, translating to 79% sensitivity and 87% specificity (95% confidence on sensitivity: 63%-92%, 95% confidence on specificity: 67%-97%, p = 0.0043 corrected, PPV = 0.88, NPV = 0.76, Matthews Correlation Coefficient = 0.65). In other words, using this approach we correctly classify 42 of 51 COVID-tested individuals who smelled Basil. One may note that selecting a 79% true positive rate still implies a 21% false negative rate, and this is potentially costly. This balance reflects a question of policy, and favoring a low false negative rate may be preferred in this pandemic^17^. With that in mind, we observe that if we select a 97% true positive rate, we incur a 27% false positive rate, reflecting 97% sensitivity and 73% specificity (95% confidence on sensitivity: 79%-100%, 95% confidence on specificity: 50%-89%, p < 0.00001 corrected, PPV = 0.63, NPV = 0.85, Matthews Correlation Coefficient = 0.342). This reflects a remarkable 3% false negative rate, well within the optimal goal of testing^17^.

**Figure 3.**
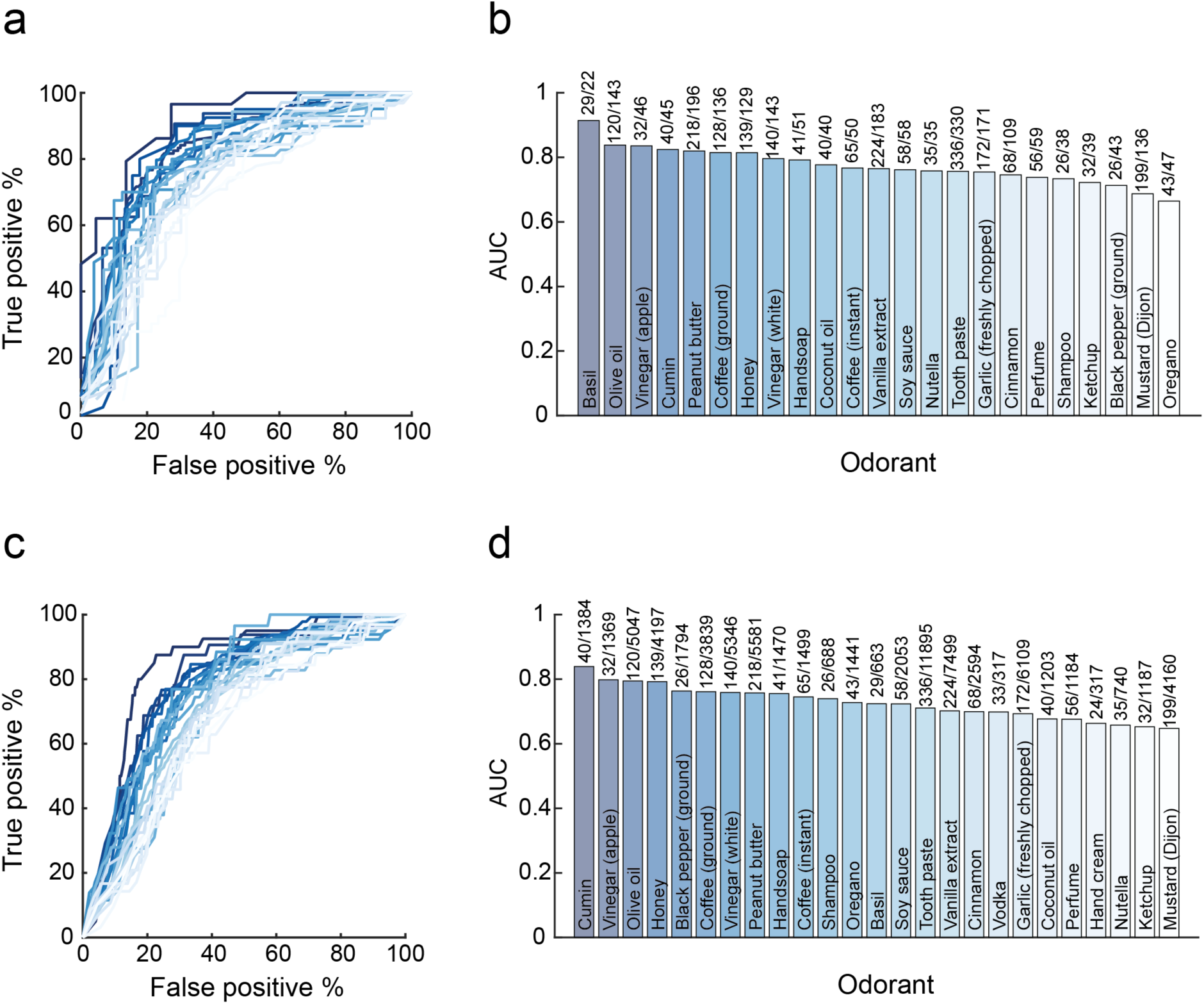
Single odorant intensity estimates indicate on COVID-19 at the individual level. **A**. ROCs based on intensity estimates of 23 odorants obtained from C19+ vs C19-participants. **B**. The AUC for each odorant, with the number of C19+/C19-participants above each bar. **C**. ROCs based on intensity estimates of 23 odorants obtained from C19+ vs C19- and C19-UD combined participants. **D**. The AUC for each odorant, with the number of C19+/C19- and C19-UD participants above each bar.

A limitation of the above result is that it relies on a significantly restricted subset of our data, namely tested individuals who also smelled Basil. To overcome this, we recalculated ROCs, now comparing between C19+ and all other participants (C19-combined with C19-UD). In other words, we assumed that untested individuals are not sick with COVID-19. Although this comparison may be weakened by unidentified C19+ individuals within the C19-UD cohort (i.e., this works against us), it allows for significantly greater sample sizes per odorant. Once again, we observe remarkable ROCs, with 4 odorants generating ROCs with AUCs greater than 0.79 (Figure 3c, 3d). The strongest, namely Cumin, had an AUC of 0.83. This implies that we could use intensity estimates of the odorant Cumin alone to identify COVID-19, and at a true positive rate of 77%, we retain a 16% false positive rate, translating to 77% sensitivity and 84% specificity (95% confidence on sensitivity: 62%-88%, 95% confidence on specificity: 50%-89%, p < 0.00001 corrected, PPV = 0.11, NPV = 0.99, Matthews Correlation Coefficient = 0.256). Although Cumin had the largest AUC in this analysis, it was rated by 1,424 participants overall, and of these only 40 participants were C19+. Similar numbers were evident in the second-best AUC, namely Apple Vinegar. However, the third best AUC, namely Olive Oil, was rated by 5,167 participants, of which 120 were C19+. This significant number of raters merits concentrating on Olive Oil as a model odorant for what this single-odorant approach might achieve. We observe that Olive Oil had an AUC of 0.79, and if we use intensity estimates of Olive Oil alone to identify COVID-19, at a true positive rate of 77%, we retain a 28% false positive rate, translating to 77% sensitivity and 72% specificity (95% confidence on sensitivity: 68%-84%, 95% confidence on specificity: 70%-73%, p < 0.00001 corrected, PPV = 0.06, NPV = 0.99, Matthews Correlation Coefficient = 0.16).

These results raise the tantalizing possibility of rapidly detecting COVID-19 by rating the perceived intensity of one odorant, such as Basil or Olive Oil, alone. Moreover, we observe that if we generate ROCs for the same 5,167 participants that smelled Olive Oil, but base them on their subjective reported symptoms (fever, cough, etc., including subjective loss in sense of smell and taste)^3^ rather than on their objective sense of smell, we obtain a ROC AUC of 0.77 (Figure 4a). Using this symptom-based ROC AUC of 0.77, at a true positive rate of 79%, we retain a 32% false positive rate, translating to 79% sensitivity and 68% specificity (95% confidence on sensitivity: 71%-85%, 95% confidence on specificity: 67%-70%, p < 0.00001 corrected, PPV = 0.05, NPV = 0.99, Matthews Correlation Coefficient = 0.15). Although this result is weaker than the olfaction-based ROC AUC of 0.79 obtained in the same individuals, this difference is not significant (StAR analysis for comparing ROCs^18^, AUC difference = 0.02, p = 0.29).

**Figure 4.**
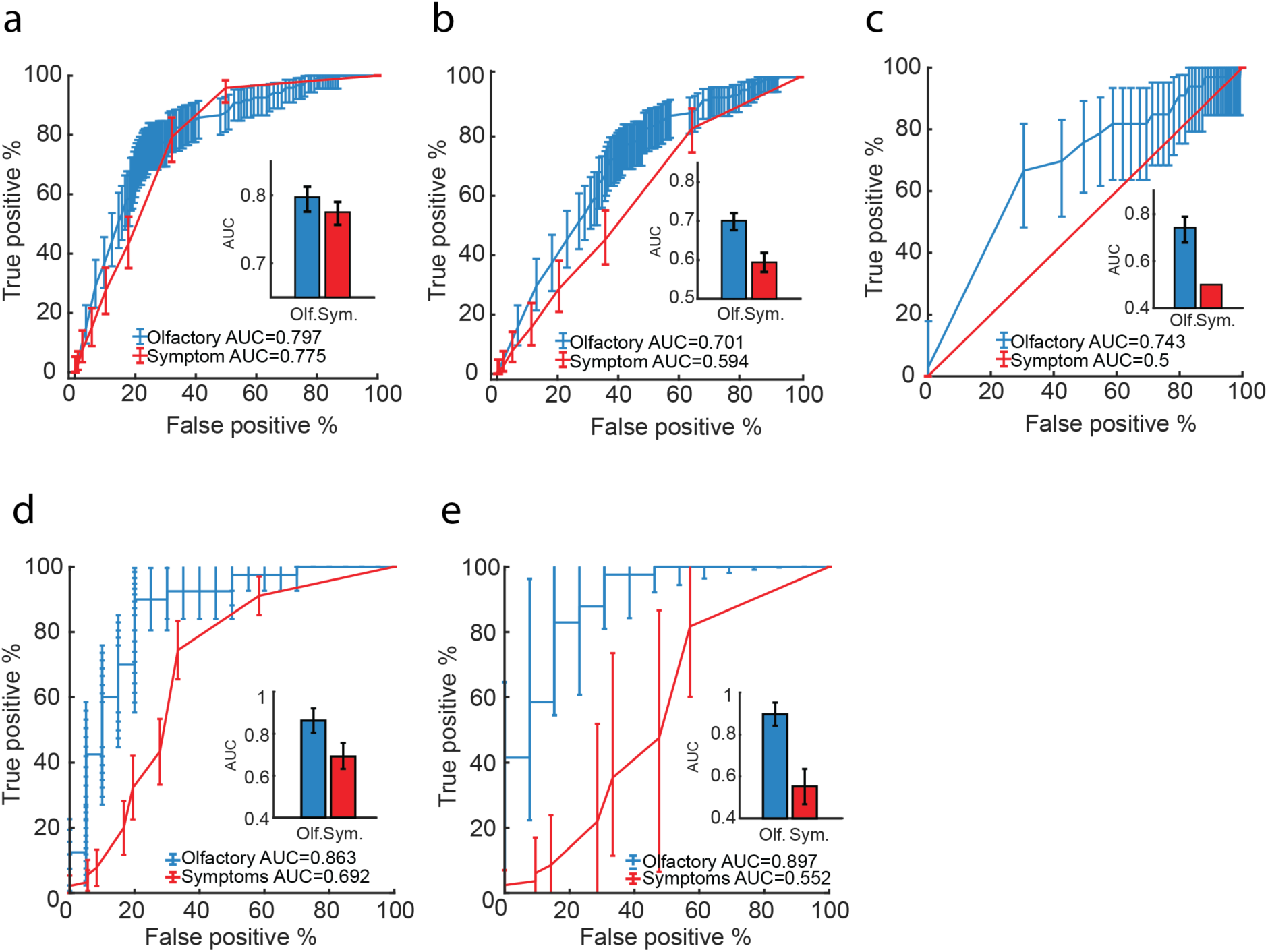
Olfactory testing is more effective than symptom checking. **A**. ROCs for all participants who smelled Olive Oil (n = 5,167 participants), based on odor intensity (blue) or reported symptoms (red). **B**. ROCs for all participants who smelled Olive Oil and had symptoms (n = 2,627 participants), based on odor intensity (blue) or reported symptoms (red). **C**. ROCs for all participants who had no symptoms (n = 7,740 participants), based on odor intensity (blue) or reported symptoms (red). **D**. ROCs based on the olfactory perceptual fingerprint (blue) or symptom reports (red) (test-set n = 60 participants). **E**. ROCs based on the olfactory perceptual fingerprint (blue) or symptom reports (red), when all participants are symptomatic (test-set n = 60 participants). Error bars on the ROCs: 95% pointwise confidence interval. Error bars for the AUC: Confidence interval derived standard deviation.

Does this minimal difference (Figure 4A) imply that single-odorant olfactory testing has no advantage over symptom checking? Although some symptom checkers have reported even stronger results than those we obtain here^19^, there are two critical points where symptom-checking alone largely fails. One such point is with individuals who all have somatic symptoms such as fever, etc., but don’t have COVID-19. Here symptom-checkers cannot avoid false positives. To address this specific point, we restricted our analysis to only participants that reported symptoms. This retained 115 C19+ symptomatic participants and 2,512 other, yet also symptomatic participants who smelled Olive Oil. Here symptoms alone gave rise to a ROC AUC of 0.59. In turn, we observe that the Olive Oil derived ROC in these same participants has an AUC of 0.70, which is significantly better (StAR analysis for comparing ROCs, AUC difference = 0.11, p = 0.00022) (Figure 4b). Thus, if we use intensity estimates of Olive Oil alone to identify COVID-19 in uniformly symptomatic populations, at a true positive rate of 75%, we retain a 40% false positive rate, translating to 75% sensitivity and 60% specificity (95% confidence on sensitivity: 66%-82%, 95% confidence on specificity: 58%-62%, p < 0.00001 corrected, PPV = 0.08, NPV = 0.98, Matthews Correlation Coefficient = 0.14).

A second point where symptom-checkers fail is at individuals who have COVID-19, but absolutely no somatic symptoms. Here symptom-checkers cannot avoid false negatives. In addressing this specific point, we are restricted by power. This is because we have only one participant who was C19+, smelled olive oil, and was completely asymptomatic. To overcome this, we use a method that allows us to compare subjects across odorants, thus retaining all 33 C19+ completely asymptomatic and 7707 other completely asymptomatic participants in our study. In brief, here we first create a combined intensity-probability measure for each participant. This measure is generated by first converting the intensity ratings of each odorant into probability scores based on the number of participants who rated the same odorant lower than the given value. The result of this step is that each participant’s 5 intensity ratings can be measured on a probability scale. Next, each participant’s 5 probability intensity scores were multiplied to produce a combined intensity probability score for each participant. This last number reflects the likelihood of the participant to rate his/her selected odorants lower than other participants. We then generated a ROC based on the participant’s combined intensity probability score. The symptoms-based ROC AUC in these participants is, by definition, 0.5, as they reported no symptoms. In turn, the odorant intensity derived ROC AUC was 0.74, a significantly better classifier (StAR analysis for comparing ROCs, AUC difference = 0.24, p < 0.0001) (Figure 4C). Thus, using odorant intensity in these participants, at a true positive rate of 67%, we retain a 30% false positive rate, translating to 67% sensitivity and 70% specificity (95% confidence on sensitivity: 48%-82%; 95% confidence on specificity: 69%-71%; p < 0.00001 corrected, PPV = 0.01 NPV = 0.99 Matthews Correlation Coefficient = 0.05). These results are not as strong as the previous results in this manuscript, but they nevertheless reflect the only remote online method we are aware of that can discriminate between completely asymptomatic C19+ and other asymptomatic individuals. In this, combined with the strong discrimination between symptomatic C19+ and other symptomatic individuals, olfactory testing with household odorants may be the most effective method currently available for remote self-testing of COVID-19.

### Olfactory Perceptual Fingerprints Outperform Symptom Checkers

Despite the undeniable attractiveness of the above simple measure, namely rating the intensity of a single odorant, we appreciate its real-world limitations. First are its susceptibility to top-down pressures and the influence of experience. Most people who would use a tool made of a single-odorant intensity estimation would understand full-well that if they report a lower intensity, they are more likely to be determined as C19+. This knowledge can influence performance in line with the results the user “wants”, whether knowingly or unknowingly. Furthermore, even without this pressure, if a person is self-testing regularly, they will very soon “know” the intensity of the odorant, and will therefore be hesitant to change their learned response. Second, several lines of evidence imply not only reduced olfactory sensitivity in COVID-19, but also altered or shifted olfactory perception^20^. Such shifts may not always be reflected in a simple intensity measure, but are reflected in a recently developed measure termed the olfactory perceptual fingerprint (OPF)^21,22^. OPFs reflect the perceptual distances between odorants within a multi-dimensional perceptual space. In other words, the measure does not ask how olive oil smells independently, or how Cumin smells independently, but rather how does Olive Oil compare to Cumin, and so on for as many odorants used to derive the measure. Thus, an OPF reflects how the world smells to an individual^21,22^. In contrast to an intensity estimate, users don’t have an intuitive sense of what goes into generating this perceptual measure, yet it remains highly informative^21,22^. In calculating OPFs for the current data we face the following tradeoff: OPFs are more powerful when they are based on a greater number of odorants. In turn, the odorants must be common across participants, so if we choose to rely on a large number of odorants, we will be left with a small number of participants. Given that a 4-odorant OPF was sufficient to categorize individuals in a manner that was related to genetic traits^21^, we stuck to this number. The 4 odorants with most participants in the current data were ‘Your toothpaste’, ‘Vanilla extract’, ‘Garlic, freshly chopped’ and ‘Peanutbutter’, which were rated by 90 C19+ participants and 36 C19-participants. A given OPF was made of all pairwise distances between these 4 odorants within the pleasantness-intensity space. We then applied an SVM classifier to the OPFs, training the classifier on one set, and testing on another. We repeated this process 500 times for different selections of training and testing sets (we assured that the testing set was made of raters who participated only once, so as to prevent any double-dipping). The symptoms-based ROC AUC in these same participants was 0.69. In turn, the OPF-derived ROC AUC was 0.86, a significantly better, and impressively strong classifier (StAR analysis for comparing ROCs, AUC difference = 0.17, p < 0.00001) (Figure 4D). In other words, unlike the single odorant (Figure 4A), the OPF provides a meaningful advantage above symptom-checking overall. Here, at a true positive rate of 90%, we retain a 20% false positive rate, translating to 90% sensitivity and 80% specificity (95% confidence on sensitivity: 80%-99%, 95% confidence on specificity: 62%-98%; p < 0.00001 corrected, PPV = 0.9, NPV = 0.8, Matthews Correlation Coefficient = 0.7). Thus, we correctly classify 52 of 60 participants in the testing set using this method.

Next, to further gauge the potential advantage of the OPF, we again restrict our analysis to participants with symptoms only. Given the double restriction of participants who all smelled the same four odorants, and all had symptoms, this analysis retained only 95 participants. Nevertheless, the symptoms-based ROC AUC in these participants was 0.55. In turn, the OPF-derived ROC AUC was 0.89, again a significantly better classifier (StAR analysis for comparing ROCs, AUC difference = 0.34, p < 0.00001) (Figure 4E). Here, at a true positive rate of 82%, we retain a 15% false positive rate, translating to 82% sensitivity and 85% specificity (95% confidence on sensitivity: 71%-93%, 95% confidence on specificity: 65%-100%; p < 0.001 corrected, PPV = 0.91, NPV = 0.70, Matthews Correlation Coefficient = 0.64). In other words, we correctly classify 50 of 60 participants in the testing set using this method. Finally, we avoid comparing symptom-checking to OPF in an all non-symptomatic cohort, because this split of the data did not retain sufficient numbers for then running separate training and testing groups.

Given that OPFs overcome two meaningful real-world limitations associated with intensity ratings alone (top-down influence, and measuring parosmia, namely shifted perception), and unlike single odorant intensity ratings, also significantly outperform symptom checking overall (Figure 4A vs. 4D), we think this should remain the olfaction-based measure of choice in reducing the findings of this study to a helpful practice.

## Discussion

There is general agreement that improved testing schemes are critical towards managing COVID-19 at the population level. However, most tests require transportation, either of the person to the test, or of the test to the person. Moreover, most testing methods then require additional transportation of the testing material, typically to a lab for processing. Such transportation is a complicating factor even in the best of times, yet now concurrent with an effort to transport vaccines, this poses even greater stress on national health systems. Moreover, once all this transportation is complete, there still remains the processing time of the test, time during which infected individuals may continue to spread the disease. Finally, beyond all this there is the matter of cost and limited availability for molecular tests. All this renders the notion of an online test incredibly appealing^23^. This need has been partially met with online symptom checkers. These have obtained some very impressive results^3,19^, yet as previously noted, they inherently all-out fail at two critical points: One is in individuals who are ill, but not with COVID-19. If an individual has a cough, fever, and a headache, symptom-checkers will most likely estimate them to have COVID-19. In turn, a person who has COVID-19 but absolutely no somatic symptoms, will go unnoticed by symptom-checkers. The latter type error is of course far more dire than the former in this case, as these people will continue to spread the disease unknowingly. Here we observe that olfactory testing, and particularly the OPF, remains effective at both these end points: correctly classifying individuals who are ill but not with COVID-19, and individuals who have COVID-19 but aren’t ill. In this respect, olfactory testing makes for a powerful and unique remote tool.

That olfaction serves as such a strong indicator of COVID-19 suggests that the olfactory impairment may be related to some fundamental aspect of this disease. Nevertheless, the mechanism by which the SARS-CoV-2 virus or the COVID-19 disease impact olfaction remains unknown^24^. The effect may be peripheral, reflecting epithelial inflamation^25^, or central, reflecting impact on the olfactory brain. Evidence for the latter can be seen in cases of COVID-19 associated olfactory pathway neuropathey^26^, and COVID-19 associated olfactory bulb edema^27^ and atrophy^28,29^. If the virus reaches the brain through olfactory pathways^30^, this is likely not via olfactory receptor neurons (ORNs), but rather through sustentacular non-neuronal supporting cells^31,32^. Like ORNs, these provide for a direct path from the intranasal periphery into the brain, and may underlie neurological aspects of the disease^33^. Moreover, given the intimate link between olfaction and respiration in the brain^34^, the olfactory path may enable the virus access to respiratory centers, thus making for a neural component in the respiratory failure associated with the disease^35^. The current study does not provide for any mechanistic insight in this respect, although the apparent difference between specific odorants in their usefulness for classification (e.g., the unique power of Basil) may reflect an avenue worthy of investigation in this respect.

This study has several limitations. First, although our overall data set is large, several of our analyses relied on restricted subsets that reduce power. Second, participants were self-selected, and this may have introduced bias. That said, we fail to identify a selection bias pattern that might underlie our effects. For example, we observe that only 4,031 participants (33.5%) reported a subjective loss of smell, so it wasn’t the case that just individuals who felt they lost their sense of smell used this tool. Third, we have no formal verification for the COVID-19 testing reported by our participants. Here too, however, any misrepresentations could have only weakened our results, as they would have only introduced added noise. Relatedly, we note that even if we had formal verification of RT-PCR tests of our participants, we nevertheless retain an upper bound on measured performance, as RT-PCR itself isn’t perfect. In other words, we observe that what we are predicting in this study is RT-PCR results, and not SARS-CoV-2 infection itself. Thus, again, our true performance level may be lower or higher than we appreciate. Finally on this front, in those diaxgnosed positive, we don’t have a time point for the diagnosis. Given that the clinical sensitivity of RT-PCR decreases with days post symptom onset, all the way down to 30% at Day 21^36^, this information is important towards characterizing the value of olfactory testing. In this respect, we also observe that given the long-term persistence of olfactory impairments in COVID-19^37^, our tool can be effective at detecting initial infection, but it is inappropriate for gauging continued infection risk from C19+ individuals.

Despite all these limitations, using a single odorant and a simple measure, namely intensity estimates, provided for a remarkably powerful tool. Although this potential speed of testing (less than 30 seconds) and simplicity of analysis are both attractive features, the applicability of this approach will be restricted to limited settings. This is primarily because of the susceptibility of this approach to interference. If a cognizant adult self-tests using a single odorant, they will easily understand the test, and may then influence it, whether knowingly or unknowingly. This limitation is overcome by the OPF. Naïve users have no intuition for this measure and how it is calculated. Moreover, the OPF is particularly sensitive to shifts in olfactory perception that do not entail a universal reduction in intensity perception alone, and such shifts may indeed be prevalent in COVID-19^20^. Finally, the OPF was significantly more effective than symptom checkers in the entire cohort. Therefore, despite this test taking longer than single-odorant rating (about 3 minutes for 4 odorants), it may be more useful. Notably, these approaches are not mutually exclusive. The on-line interaction can remain the same, and the analysis for a one-time user can favor odorant intensity, yet the display and analysis of repeated users can rely on OPFs. Such repeated tests may gain added power^38^, and provide the basis of a testing-regimen, a critical aspect of population-level curtailment^1^. We think this is a rare case where something so utterly simple may prove so utterly valuable.

## Supporting information

raw data

## Data Availability

Raw data as well as matlab files are attached.
code for processing creating the figures is available.

https://gitlab.com/snitz/smelltracker_article

## Acknowledgments

This study was supported by a Weizmann CoronaVirus Emergency Fund grant from Miel de Botton and a Weizmann CoronaVirus Emergency Fund grant from Sonia T. Marschak.

## Author contributions

Developed concepts: All authors.

Built web-tool: KS; DH; NH; RW.

Translated to native language: RW; EM; SK; AM; BI; AA; GE; MO; CP; NB; ZM; ERM; CD; SAS;

FMS; CF; MS; KT; MB; TH; JNL

Analyzed data: KS; RW; AR; EM

Figures and visualization: KS; RW; AR; EM; OP

Wrote first draft of paper: KS; RW; AR; EM; NS

Edited final draft: All authors

## METHODS

### Data availability

All raw data are available for download in Supplementary Data File 1

### Code availability

All code used in this manuscript will be available for download at https://gitlab.com/snitz/smelltracker_article

### Recruitment

There was no systematic recruitment. Each participating lab tried to inform the local media in their country of residence. This resulted is several news stories published in several countries, and these led to dissemination. Participants were directed to the web-tool at www.smelltracker.org where they participated anonymously in an interaction that was approved by the Weizmann Institute of Science IRB committee, and by the Wolfson Hospital Helsinki Committee. We note that during the reported time period we had 12800 participants, but of these 780 participants reported only partial data, retaining only 12,020 participants for full analysis.

### Web-tool

The tool was written in open-source Drupal 8 (drupal.org). On it, participants first created a unique login to facilitate repeated testing. Next, participants provided details regarding age, sex (Woman/Man), and country of residence (here we made a mistake in that the country pull-down menu did not start from an empty space, but rather from “India”. Thus, participants who failed to answer this question were registered as from India by default. For this reason, we are unable to faithfully include India in the country-specific analyses (Figure 2)). Next, participants selected five odorants to rate. We opted for five odorants, rather than a larger number, to strike a balance between increased reliability, where more assessments render more reliable data^39^, and low burden of participation. Each odorant was selected from a separate category with a fixed list of common household odorants (Supplementary Table 1). This list was generated in coordination with the participating labs in order to assure cultural diversity. Two odorant categories contained odorants with reduced trigeminal components (e.g., vanilla extract), and three categories had increased trigeminal components (e.g., vinegar). Participants made their odorant selections upon first use of the tool, and were then automatically prompted to use the same odorants on subsequent uses. Participants then proceeded to smell each odorant and rate its perceived intensity and pleasantness on separate visual analog scales, one for intensity ranging from very weak to very strong, and the other for pleasantness ranging from very pleasant to very unpleasant. These scales were coded in the system as ranging from 0 to 100. Participants could smell the odorant as often as they liked, and there was no time limit applied. Following the ratings, participants were asked whether they had been tested for COVID-19 (No; Yes-Pending; Yes-Positive; Yes-Negative), and whether they are currently experiencing any COVID-19 symptoms (Fever; Cough; Shortness of breath or difficulty breathing; Tiredness; Aches; Runny nose; Sore throat; Loss of the sense of smell; Loss of taste; No symptoms). Finally, participants were presented with a graph depicting their olfactory perceptual fingerprint as it related to the average scoring, and if they participated again, the graph depicted the evolution of their perception over time. In addition to the graph, participants were presented with a text informing them whether their perception was within range of our participants, or aberrant. That said, at this stage the text did not explicitly inform participants that we predict them to be C19+, as we did not yet receive regulatory permission to make such a statement.

### Statistical analyses

All analyses were conducted using Matlab software, and the complete data file allowing full recreation of these results is in Supplementary Data File 1.

For initial analysis of intensity and pleasantness, we restricted our analysis to 23 odorants that had more than 25 C19+ raters. This gave rise to 46 distributions of ratings, of which only 18 and 33 for intensity and pleasantness respectively, were normally distributed. Given non-normal distributions, we applied a two-sided Kolmogorov-Smirnov test to all C19+ and C19-intensity and pleasantness comparisons. In the individual odorant follow-up comparisons we estimated effect size using the Eta squared effect size measure^40^.

### Country-specific correlations

between odorant ratings and rates of COVID-19 were calculated as follows: To produces rates of COVID-19 time-series, we conducted 2 steps: 1. The number of daily cases in each country was obtained from the Johns Hopkins Coronavirus Resource Center^12^. 2. We calculated a 7-day moving average for the dates between March 15, 2020 and September 30, 2020. For national intensity ratings time-series, we conducted 3 steps: 1. Average intensity ratings of the 5 odorants were calculated for each entry. 2. Mean intensity ratings were inverted by subtracting them from 100. This was done so that higher values imply greater smell loss. 3. A 5-day moving average was calculated by averaging all ratings in the span of 7 days. We used this moving average to match the cases span. After obtaining these two values, a cross-correlation between the daily ratings and inverse intensity was then calculated (using the xcorr function in Matlab). The cross-correlation analysis resulted in a correlation between the two signals for different lags (between 14-days earlier to 14-days later response) in the inverse intensity signal. The lag that produced the maximal correlation between the two signals was chosen for the analysis. The Pearson correlation between daily cases and lagged inverse intensity was calculated. Daily cases time-series, inverse intensity signal and lagged inverse intensity signal are shown in figure 2.

### ROCs

were calculated using standard technique^41^. We used a moving cutoff point on a continuous scale and at each point measured the true positive (TPR) and false positive (FPR) ratios which result from selecting that cutoff. All confidence intervals in ROC plots were calculated using a 1000 iteration bootstrapping of the scores. To compare between ROCs, we used a non-parametric test based on the AUC of the curves^18^.

### Olfactory Perceptual Fingerprints

were calculated as previously described in detail^21,22^. In brief, we selected the 4 odorants which were rated most often. Those were ‘Your toothpaste’, ‘Vanilla extract’, ‘Garlic, freshly chopped’ and ‘Vinegar white’. This choice restricted our analysis to 3274 participants. We then calculated an individual *olfactory fingerprint* by computing all the pairwise distances (i.e. derived similarity) between these 4 odorants. For a measure of distance between odorant k and odorant m we used the following equation:

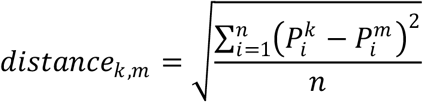

Where 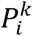 is the perceptual rating of odorant *k* using descriptor *i*. and 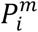 is the perceptual rating of odorant *m* using descriptor *i*. In words: the distance between odorant *k* and odorant *m* is the square root of the mean of the squared difference between the two perceptual ratings (intensity and pleasantness) for those two odorants. Once all the pairwise distances are computed we can notate an *olfactory fingerprint* in the following form:

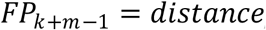

Where k = 1…N and m = k+1…N. In words; each element of the *olfactory fingerprint* is the distance between pairs of odorants, where each pairwise distance is calculated only once (since *distance*_*k,m*_ = *distance*_*m,k*_) and the distance between an odorant to itself is omitted (since *distance*_*k,k*_ = 0). Consequently, if we here use 4 odorants to construct an *olfactory fingerprint* the resulting olfactory fingerprint will have 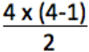, or in other words 6, elements.

## Legends

**Extended Data Table 1**

The list of odorants participants could select from. Participants were instructed to select one odorant from each of the 5 categories, summing at 5 odorants for rating of intensity and pleasantness.

**Supplementary Data File 1**

This file contains all the manuscript raw data

**Extended Data Figure 1.**
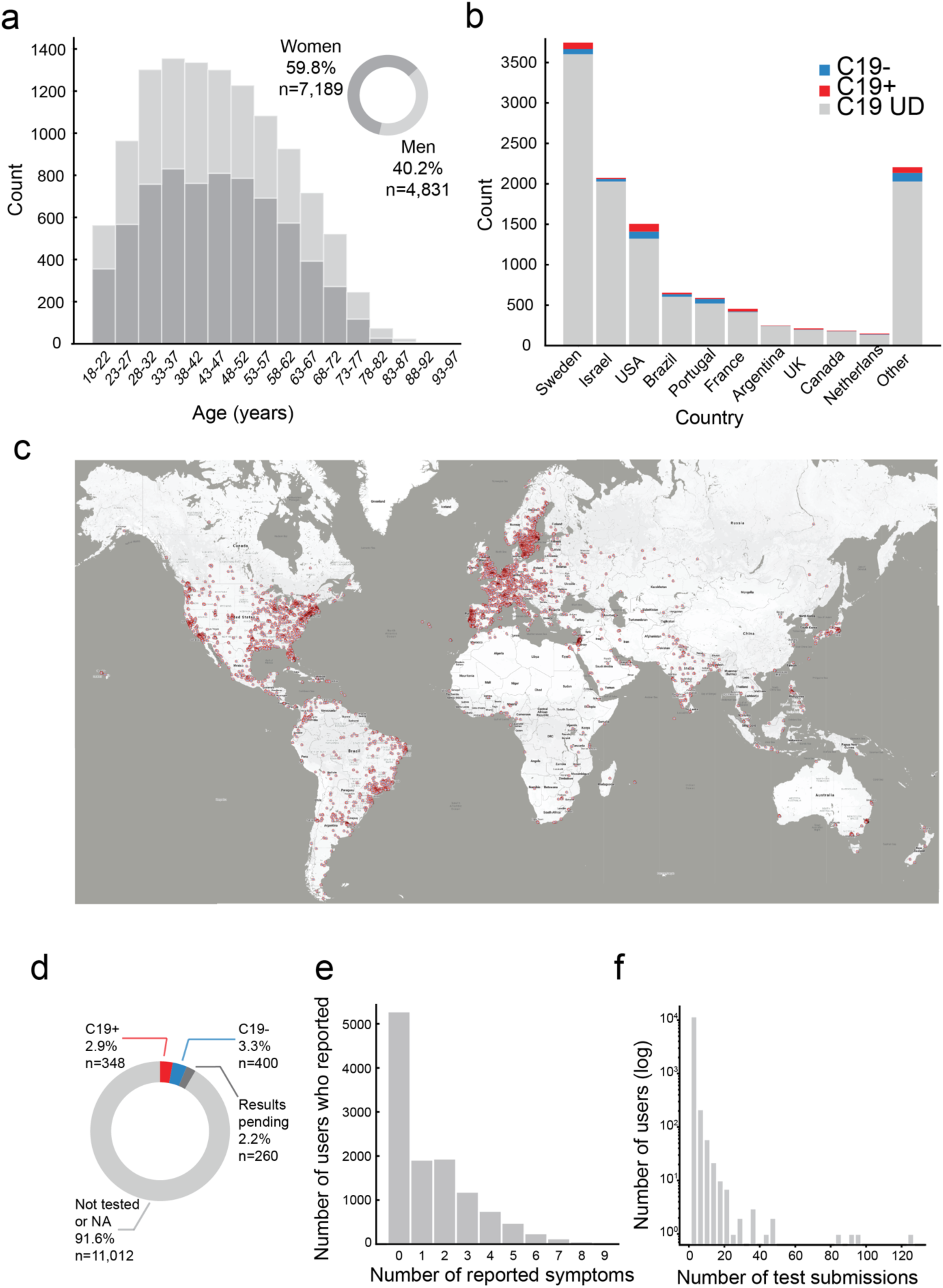
Characterization of 12,020 participants. **A**. Age and gender distribution of participants. **B**. Number of participants and their COVID-19 status from the 10 highest-participation countries (see comment on India in Methods). **C**. Geographical distribution of respondents, each dot is a participant, overlapping dots not shown to maintain clarity. **D**. The distribution of C19+, C19-, and C19-UD in the sample. **E**. Distribution of number of somatic symptoms reported by participants. **F**. Distribution of number of submissions per participant.

