## extended data table for "A Novel Olfactory Self-Test Effectively Screens for COVID-19"

**Extended Data Table 1**

| **Category 1** | **Category 2** | **Category 3** | **Category 4** | **Category 5** |
|---|---|---|---|---|
| Vanilla extract | Peanut butter | Mustard (Dijon) | Garlic (freshly chopped) | Your toothpaste |
| Nutella | Coconut oil | Vinegar (white) | Camembert cheese | Your hand soap |
| Honey | Olive oil | Horseradish (jar) | Canned tune | Your laundry detergent |
| Strawberry jam | Basil | Wasabi | Blue cheese | Your shampoo |
| Apricot jam | Oregano | Onion (freshly chopped) | Canned sardines | Your hand cream |
| Apple juice (not fresh) | Parsley | Vinegar (apple) | Mushrooms | Your body lotion |
| Orange juice (not fresh) | Cilantro | Black pepper (ground) | Boiled egg | Your perfume |
| Lemonade (not fresh) | Dill | Menthol gum | Pickled herring | Your hand sanitizer |
| Peach nectar (not fresh) | Cardamom | Mint (fresh) | Cumin | Your sunscreen |
| Pear nectar (not fresh) | Thyme | Mint (gum) | Soy sauce | Your baby oil |
| Grapefruit juice (not fresh) | Nutmeg | Mint (tea) | Sauerkraut | |
| Pineapple juice (not fresh) | Caraway | Sesame oil | Coffee (ground) | |
| Banana nectar (not fresh) | Bay leaves | Vodka | Coffee (instant) | |
| Cinnamon | Ketchup | Clove | Tea (black) | |
| Maple syrup | | Vinegar (balsamic) | Tea (earl gray) | |
| | | Vinegar (red) | | |
| | | Mustard (ordinary) | | |
